# Transcranial direct current stimulation combined with alcohol cue inhibitory control training reduces the risk of early alcohol relapse: a randomized placebo-controlled clinical trial

**DOI:** 10.1101/2021.06.24.21259446

**Authors:** Macha Dubuson, Charles Kornreich, Anne Vanderhasselt, Chris Baeken, Florent Wyckmans, Clémence Dousset, Catherine Hanak, Johannes Veeser, Salvatore Campanella, Armand Chatard, Nemat Jaafari, Xavier Noël

## Abstract

**Background:** Approximately half the people with alcohol use disorder (AUD) relapse into alcohol reuse in the few weeks following withdrawal treatment. Brain stimulation and cognitive training represent recent forms of complementary interventions in the context of AUD.

**Objective:** To evaluate the clinical efficacy of transcranial direct current stimulation (tDCS) over the dorsolateral prefrontal cortex (DLPFC) combined with alcohol cue inhibitory control training (ICT) as part of rehabilitation.

**Methods:** A randomized clinical trial was conducted on patients (n=125) withsevere AUD at a withdrawal treatment unit. Each patient was randomly assigned to one of four conditions, in a 2 [verum vs. sham tDCS] x 2 [alcohol cue vs. neutral ICT] factorial design. The primary outcome of the treatment was the measured abstinence rate after two weeks or more (up to one year).

**Results:** Verum tDCS improved the abstinence rate at the 2-week follow-up compared to the sham condition, independently of the training condition (79.7% [95% CI = 69.8-89.6] vs. 60.7% [95% CI = 48.3-73.1]; p = 0.02). A priori contrasts analyses revealed higher abstinence rates for the verum tDCS associated with alcohol cue ICT (86.1% [31/36; 95% CI= 74.6-97.6]) than for the other three conditions (64% [57/89; 95% CI = 54-74]). These positive clinical effects on abstinence did not persist beyond two weeks after the intervention.

**Conclusions:** AUD patients who received tDCS applied to DLPFC showed a significantly higher abstinence rate during the weeks following rehabilitation. When combined with alcohol-specific ICT, brain stimulation may provide better clinical outcomes.

**Trial Registration:** ClinicalTrials.gov number NCT03447054

## Introduction

With around 283 million people aged 15 years and older facing severe problems related to alcohol use, alcohol use disorder (AUD) continues to represent a substantial public health problem around the world [1]. In the most severe cases (which require physical follow-up), patients with severe AUD require hospital weaning assisted by medication [2]. However, a large majority of these patients, even if properly detoxified, are at risk of relapse within weeks of hospital discharge [3–5].

According to several neurocognitive models, addiction is difficult to overcome due to several brain mechanisms and related cognitive and emotional processes that are resistant to changes in reinforcement contingency (e.g., devaluation) [6]. The mechanisms involved in this state of flexibility, reflecting the so-called “addiction paradox”, are sensitized automatic processes operating with little intention, awareness, and effort [7]. Additionally, the effortful supervisory system which includes a variety of cognitive functions (e.g., response inhibition and mental shifting) aimed at modulating spontaneous responses (e.g., suppression), is dramatically affected in individuals with AUD. This dual-process approach of behavior and choice [8,9] applied to addiction [7,10–12] has led to the idea that, to be effective, a clinical intervention should modify not only deliberative processes (e.g., reappraisal, inhibitory control) but also cognitive, behavioral, and automatic emotional responses.

Inhibitory Control Training (ICT) [4,13–15] and neuromodulation [16,17] are non-pharmacological interventions that have been used to improve clinical trajectories in participants with unhealthy behaviors [4,13–15], including AUD [18,19]. For instance, to strengthen alcohol/response inhibition associations, heavy alcohol drinkers were trained to associate alcohol-related pictures with a No-Go (not-pressing) response and non-alcohol cues with a Go (pressing) response [14,20]. However, meta-analyses showed a small-to medium-size effect on reducing alcohol consumption and relapse rate, which raised the issue of their clinical utility [21,22]. Meanwhile, transcranial direct current stimulation (tDCS) has been shown to be a tolerable, safe, easy-to-apply, and cost-effective alternative non-pharmacological treatment for addiction [23,24]. The dorsolateral region of the prefrontal cortex (DLPFC) has been the primary target for various clinical purposes [23], including reduced craving and alcohol use reduction [19,25,26]. Although promising, the lack of a multisession setup combined with simultaneous behavioral and cognitive efforts might limit the clinical impact of tDCS in psychiatry [26–29]. Indeed, the effects of tDCS may be mainly state-dependent; in other words, there exists an interaction between external stimuli and the underlying state of the stimulated region or network [30]. In the same vein, we refer to the ‘activity-selectivity’ hypothesis, which states that tDCS preferentially modulates active over inactive neural populations [31]. When used in monotherapy, tDCS might primarily target deliberate processes, thus leaving automatic processes unchanged [18].

Consequently, tDCS and ICT in combination, can be used in response to alcohol-related cues to protect people against alcohol relapse by modifying the suboptimal interaction between the strong alcohol-related response and the weakened control over this response [7,10–12,32]. Indeed, when prefrontal regions are stimulated, tDCS is assumed to slightly improve several functions related to executive functioning [19,33–36]. Additionally, systematically matching a No-Go response with motivational content reduces its positive valence [37,38]. Therefore, when combined, these interventions could provide the greatest clinical benefits in AUD.

To improve the clinical trajectories of patients with AUD, we combined a multisession alcohol cue ICT with stimulation of the DLPFC with tDCS. To date, only two clinical trials in patients with AUD have combined tDCS with cognitive alcohol bias modification, with little evidence of positive changes in the clinical trajectory of patients with AUD during the year following the intervention [39,40]. However, both studies applied four sessions of tDCS with the anode located at F3 and the cathode at F4 combined with a behavioral paradigm (approach-avoidance task) unrelated to inhibitory control, while recommendations pointed to an alternative montage (anode-F4 and cathode-F3 repeated for at least five sessions) [26,28].

By following this recommendation, we tested the hypothesis that the combination of tDCS and alcohol cue ICT would reduce the risk of relapse in patients with severe AUD more than other interventions using sham tDCS and neutral ICT. It should be noted that although the vast majority of clinical trials in inpatients have focused on behavioral interventions and brain stimulation that reduce alcohol relapse after three months or more, a large proportion of patients relapse within a couple of weeks following discharge [3–5]. Therefore, we primarily focused on early alcohol relapse, that is, the two weeks after discharge. We also investigated whether this reduction was still present several months after the end of the detoxification treatment (up to one year) and whether this clinical effect was mediated by several psychological mechanisms (i.e. cognitive control, craving, mood and cognitive bias).

## Material and methods

### Participants

Patients were recruited while undergoing alcohol rehabilitation at the Brugmann University Hospital in Brussels, Belgium. Inclusion criteria included French-speakers between 18 and 65 years of age with severe AUD requiring alcohol rehabilitation and the desire to stay sober for at least the first six months after detoxification. The exclusion criteria based on th**e** International Neuropsychiatric Interview [41] included neurological history (epilepsy, head injury, and stroke), mental confusion or severe cognitive impairment, schizophrenia, chronic psychotic disorders, bipolar type 1 disorder, metal in the brain, and pregnancy.

### Design

The study was a single-blind (participants) and parallel 2 [verum vs. sham tDCS] x 2 [alcohol cue vs. neutral ICT] full-factorial design. Patients were randomized using a computer software program that generates the random sequence to one of the four experimental conditions: (1) verum tDCS during alcohol cue ICT (AICT); (2) verum tDCS and Neutral ICT (NICT); (3) sham tDCS and AICT; and (4) sham tDCS and NICT. Recruitment was conducted from January 2018 to March 2021. Figure 1 depicts the screening and recruitment information.

**Figure 1.**
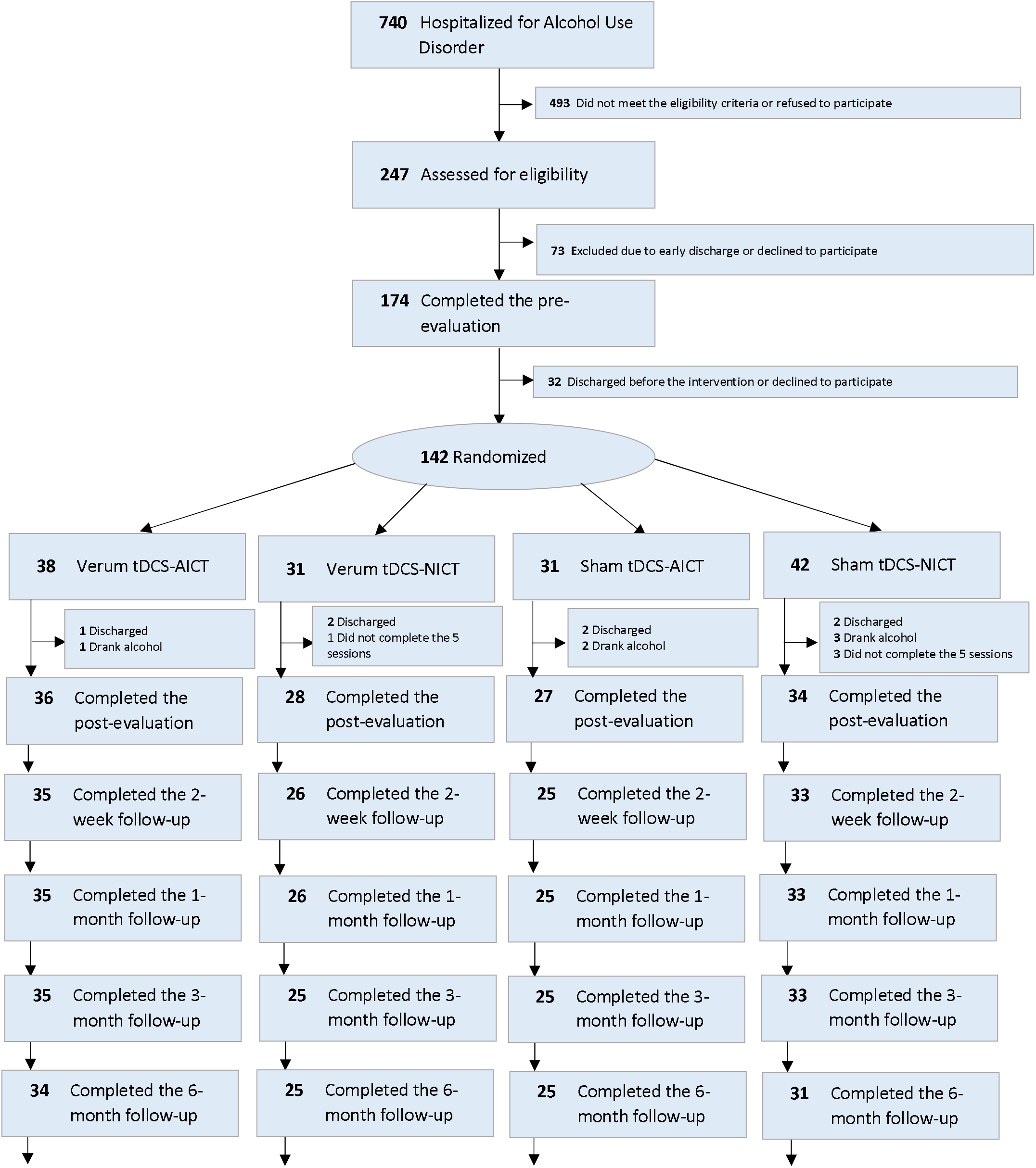
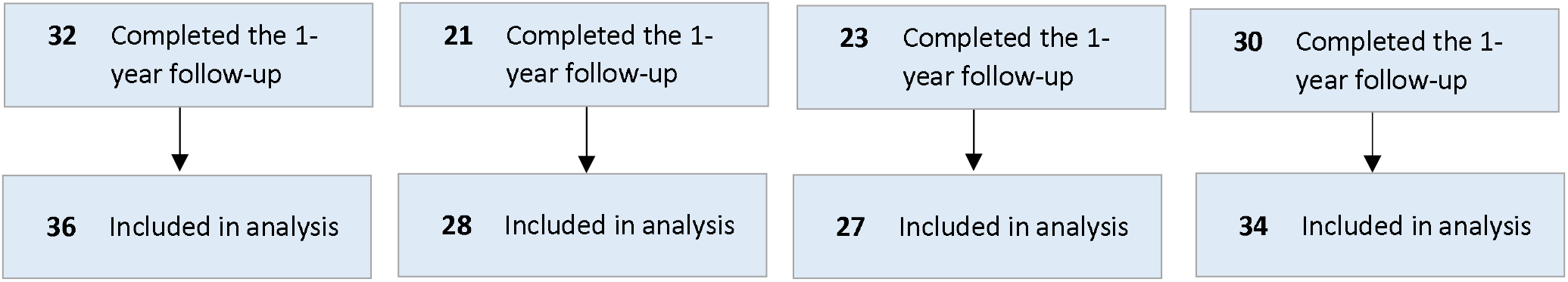
Screening, recruitment, randomization, treatment completion, and follow-up completion data. Abbreviations: AICT, Alcohol Cue Inhibitory Control Training; NICT, Neutral Inhibitory Control Training.

The intervention consisted of five consecutive daily 20-minute sessions of a simultaneous combination of tDCS and ICT (Monday to Friday). The ICT was started immediately after the tDCS was put in place, and the instructions were correctly understood. At baseline (the Friday before the intervention) and after the intervention (the Monday after the intervention), the measurements of working memory, craving, response inhibition, verbal fluency of alcohol words and mood were recorded. The order of cognitive tasks was administered at random, followed by clinical assessment.

### Transcranial direct current stimulation protocol

A bilateral direct current of 2 mA with a 30 s ramp-up and 30 s ramp-down was applied for 20 min on the DLFPC. This direct current was delivered with a 1×1 low-intensity transcranial DC stimulator linked to two electrodes, with a surface sponge of 35 cm^2^. The anode was placed over F4 (right DLFPC) and the cathode over F3 (left DLFPC), according to the 10-20 international system for electroencephalogram electrode placement (vertical placement). In the sham procedure, the electrodes were placed in the same positions but without stimulation, and only ramp-up and ramp-down were applied [42]. The intervention included five consecutive 20-minute daily sessions that simultaneously combined tDCS and ICT.

### Inhibitory control training protocol

Concurrently with tDCS, patients underwent alcohol cue or neutral ICT. In a modified version of a Go/No-Go task presented on a 15-inch laptop, participants were instructed to press a response key whenever a letter (P or R) was displayed in one of the four corners of the picture with a probability of .5, and not to respond when an alternative letter was displayed. The No-Go signal always matched the alcohol pictures, and the Go signal always matched the sports pictures. The neutral picture was associated with the Go signal, with a probability of .5. The session began with a five-second countdown, after which a fixation cross appeared for 1500 ms-2500 ms at random, followed by an image appearing alone for 500 ms. The Go or No-Go signal was displayed in one of the four corners of the picture for 2000 ms. The task included five blocks of 64 trials: 32 Go and 32 No-Go; the same 32 images repeated twice for the AICT (eight alcohol, eight sport, and 16 neutral images), and the same 16 neutral images four times for the NICT. The participants had 2000 ms to give a response before they received one of three possible feedbacks: “Too late,” “Correct,” or “False” (**Figure 2**). The tasks, programmed with E-Prime 3.0, consisted of 360 trials and lasted approximately 20 min.

### Primary outcomes

All patients received a follow-up phone call based on the Timeline Follow-Back method [43] at 2 weeks and 1, 3, 6, and 12 months after discharge from the hospital. They were asked about their alcohol use (i.e., the amount of alcohol consumption at various levels, average number of drinks per day consumed, and maximum number of drinks consumed each day) with relapse defined as consumption of 60 g of alcohol on any occasion, on a single day. It should be mentioned that all patients with AUD reported aiming to remain sober for a minimum of 6 months after rehabilitation. In addition, during the phone call, patients who reported consuming at least 60 g of alcohol also showed loss of control over alcohol, as evidenced by a “no” response to “Did you intend to drink as much as you did?”. To increase the accuracy of the information, we contacted a person close to the patient via telephone (e.g., their general practitioner or a family member) who fully confirmed that the return to alcohol use was associated with guilt, a phenomenon deemed to be due to violation of the personal goal of abstaining from alcohol. Finally, a certified psychologist in charge of clinical follow-up reported that all the patients who resumed drinking alcohol within two weeks of their discharge presented a persistent harmful alcohol use pattern, which required additional clinical counseling and re-treatment over the next 3-9 months.

### Secondary outcomes

Secondary outcomes referred to the psychological mechanisms involved in reducing relapse into alcohol use. We examined several parameters measuring executive control, mood and affect and positive/negative alcohol associations on two separate occasions (i.e., three days before and three days after the intervention). The craving for alcohol was measured immediately before and after each intervention.

1. The craving was scored by a *visual analog scale (VAS)* including three single-items: (A) simple question (‘*I want to drink alcohol*.’); (B) craving induced by mental images (‘*Please think about alcohol, visualize it, imagine how it tastes and smells, does that make you want to drink?*’); (C) craving induced by a photo of the alcohol section of a store (‘*Please look at the picture below, does it make you want to drink?’*) (Range, 0-10 for each dimension).
2. The positive and negative affects were measured using the *Beck Depression Inventory II (BDI-II)* [44], which assessed the severity of depressive symptoms (21 items; range, 0-63), and the *Positive Affect and Negative Affect Score (PANAS)* [45] evaluating the positive and negative affect in two scores (both 10 items; range, 10-50).
3. The positive/negative associations toward alcohol and sport were measured by the alcohol verbal fluency task (adapted from [46]). The patients were required to generate in one minute as many words as possible related to either *alcohol* or *sports*. The order of these tasks was counterbalanced between subjects, and we kept both measures (T1 and T2). Words were scored by three experimenters as either *positive, negative*, or *neutral*. Inter-rater reliability was assessed using two-way mixed, absolute agreement, average measures inter-class correlation coefficients. ICC= 0.86 for positive, 0.98 for negative and 0.96 for neutral words. Additionally, participants carried out a semantic verbal fluency task consisting of generating a maximum number of names of animals within a one-minute time frame.
4. The motor inhibition performance was measured using the *Stop Signal Task (Stop-it)* [47]. The participant was required to press a button every time a “Go” signal was displayed (square or circle in the screen in 75% of cases). They were also required not to press the button when a random auditory “stop signal” was played (in 25% of cases). The subject was thus required to use motor inhibition to stop the motion to press the button. The *horse-race model* [48] proposed the *Stop-Signal Reaction Time (SSRT)*, which estimates stop signal latency. A longer SSRT indicated lower inhibitory control.
5. The working memory performance was measured using the *Operation Span task (OSPAN)*. A series of words appeared one by one on a screen (2 to 6 words, e.g., ‘blanket, ‘grass’) and between each word presentation, a mental calculation problem appeared (e.g., ‘(6/2)-1=4?’). The participants were instructed to memorize the series of words while responding to the mental calculation problem appearing between each word. The partial credit unit (PCU) score was the sum of the percentages of correct words in a series of words [49]. The higher the PCU, the better the subject performed on the working memory task.

### Clinical evaluation

The *Alcohol Use Disorder Identification Test (AUDIT*) [50] was used to score the severity of alcohol problems (range, 0-40). The *Craving Experience Questionnaire (CEQ)* [51] scored the intensity and frequency of craving in the previous week (range, 11-77). The *Impulsivity Behavior Scale (UPPS-P)* [52] assessed five facets of impulsivity (positive urgency, negative urgency, lack of premeditation, lack of perseverance, and sensation seeking; 20 items; range, 20-80). The *State-Trait Anxiety Inventory (STAI-Y)* [53] was also administered (both 20 items; range, 20-80).

### Statistical analyses

The demographic variables and questionnaires at baseline were compared between the conditions using a one-way analysis of variance (ANOVA) and the Pearson chi-square test. A binomial test was performed to compare the percentage of correct assumptions of being part of the verum tDCS intervention.

The binary outcome (relapse vs abstinence) was analyzed using logistic regressions with a priori contrasts. To test the effects of verum tDCS vs sham tDCS and AICT vs NICT on abstinence rate, we first used logistic regression with a simple orthogonal contrast evaluating the main tDCS effect (C1, verum vs. sham), the main ICT effect (C2, neutral vs. alcohol cue), and the interaction of these contrasts (C3) on the abstinence rate. Second, a deviation contrast was used to test the superiority of the verum tDCS-AICT condition effect on abstinence rate more specifically, compared to all the other conditions. The exact matrices used are reported in Table 2.

Repeated and mixed ANOVAs (Conditions x Time) were performed to assess the main effect of the intervention on craving (VAS), depressive symptoms (BDI) and affect (PANAS), verbal fluencies

(alcohol, sport, neutral), inhibition response (SSRT), and working memory (OSPAN). All analyses were performed with IBM SPSS statistics (v. 26), except for the logistic regressions, which were performed on R Studio (v. 1.4.1103). The threshold for significant effect was p < .05.

In the supplemental material, we reported the results at 1-, 3-, 6-, and 12-month follow-ups (see supplemental eFigure1 and eTable1), the ICT data analysis (see supplemental eTable2), self-scoring of ICT images (see supplemental eTable3), and the patient’s appreciation of the treatment (see supplemental eResults).

## Results

### Demographic variables

The clinical trial was offered to 756 hospitalized patients between February 2018 and March 2020. A total of 247 patients agreed to participate in the initial clinical interview, and 174 patients started the experiment (T1). Seventy-three patients were excluded because they met the exclusion criteria (e.g., epilepsy, bipolarity) or had decided not to participate for various reasons (for example, too much time investment, low motivation for the clinical trial, or fear of tDCS). Thirty-two patients discontinued their participation after T1 due to premature discharge. A total of 142 patients completed the five intervention sessions (see Figure 1).

Seventeen participants missed the post-intervention assessment due to alcohol consumption or early discharge. The final analysis included 125 participants.

Table 1 presents the clinical and sociodemographic characteristics of the participants. The patients received a detoxification regimen consisting mainly of decreasing doses of diazepam. There was no significant difference in the initial dose of diazepam administered between participants in the four conditions (F(3, 122)=1.505, p=.217).

**Table 1.**
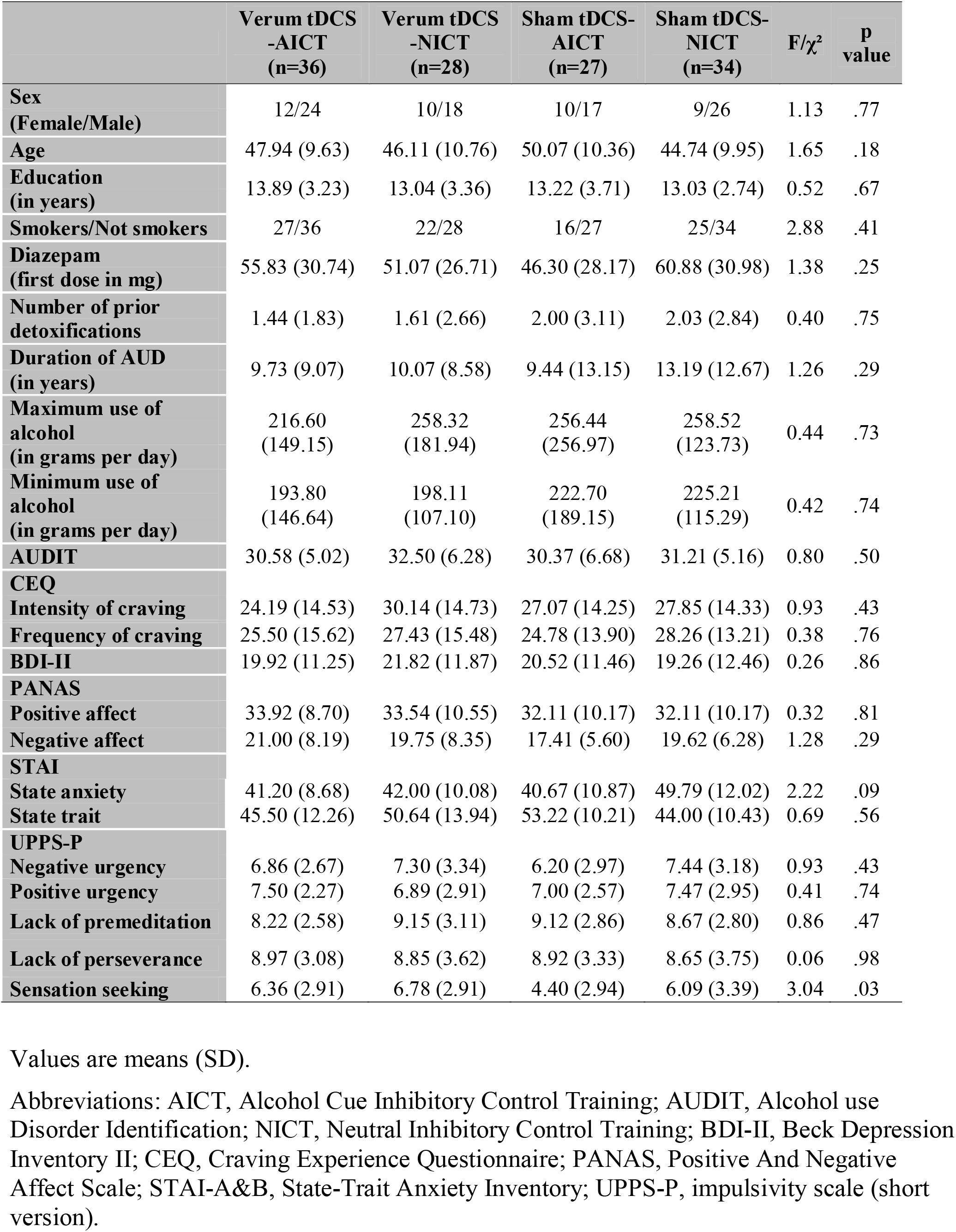
Demographic and clinical characteristics of the sample at baseline.

|  | **Verum tDCS**<br>**-AICT**<br>**(n=36)** | **Verum tDCS**<br>**-NICT**<br>**(n=28)** | **Sham tDCS-**<br>**AICT**<br>**(n=27)** | **Sham tDCS-**<br>**NICT**<br>**(n=34)** | **F/$\chi^2$** | **P**<br>**value** |
| --- | --- | --- | --- | --- | --- | --- |
| **Sex**<br>**(Female/Male)** | 12/24 | 10/18 | 10/17 | 9/26 | 1.13 | .77 |
| **Age** | 47.94 (9.63) | 46.11 (10.76) | 50.07 (10.36) | 44.74 (9.95) | 1.65 | .18 |
| **Education**<br>**(in years)** | 13.89 (3.23) | 13.04 (3.36) | 13.22 (3.71) | 13.03 (2.74) | 0.52 | .67 |
| **Smokers/Not smokers** | 27/36 | 22/28 | 16/27 | 25/34 | 2.88 | .41 |
| **Diazepam**<br>**(first dose in mg)** | 55.83 (30.74) | 51.07 (26.71) | 46.30 (28.17) | 60.88 (30.98) | 1.38 | .25 |
| **Number of prior**<br>**detoxifications** | 1.44 (1.83) | 1.61 (2.66) | 2.00 (3.11) | 2.03 (2.84) | 0.40 | .75 |
| **Duration of AUD**<br>**(in years)** | 9.73 (9.07) | 10.07 (8.58) | 9.44 (13.15) | 13.19 (12.67) | 1.26 | .29 |
| **Maximum use of**<br>**alcohol**<br>**(in grams per day)** | 216.60<br>(149.15) | 258.32<br>(181.94) | 256.44<br>(256.97) | 258.52<br>(123.73) | 0.44 | .73 |
| **Minimum use of**<br>**alcohol**<br>**(in grams per day)** | 193.80<br>(146.64) | 198.11<br>(107.10) | 222.70<br>(189.15) | 225.21<br>(115.29) | 0.42 | .74 |
| **AUDIT** | 30.58 (5.02) | 32.50 (6.28) | 30.37 (6.68) | 31.21 (5.16) | 0.80 | .50 |
| **CEQ** |  |  |  |  |  |  |
| **Intensity of craving** | 24.19 (14.53) | 30.14 (14.73) | 27.07 (14.25) | 27.85 (14.33) | 0.93 | .43 |
| **Frequency of craving** | 25.50 (15.62) | 27.43 (15.48) | 24.78 (13.90) | 28.26 (13.21) | 0.38 | .76 |
| **BDI-II** | 19.92 (11.25) | 21.82 (11.87) | 20.52 (11.46) | 19.26 (12.46) | 0.26 | .86 |
| **PANAS** |  |  |  |  |  |  |
| **Positive affect** | 33.92 (8.70) | 33.54 (10.55) | 32.11 (10.17) | 32.11 (10.17) | 0.32 | .81 |
| **Negative affect** | 21.00 (8.19) | 19.75 (8.35) | 17.41 (5.60) | 19.62 (6.28) | 1.28 | .29 |
| **STAI** |  |  |  |  |  |  |
| **State anxiety** | 41.20 (8.68) | 42.00 (10.08) | 40.67 (10.87) | 49.79 (12.02) | 2.22 | .09 |
| **State trait** | 45.50 (12.26) | 50.64 (13.94) | 53.22 (10.21) | 44.00 (10.43) | 0.69 | .56 |
| **UPPS-P** |  |  |  |  |  |  |
| **Negative urgency** | 6.86 (2.67) | 7.30 (3.34) | 6.20 (2.97) | 7.44 (3.18) | 0.93 | .43 |
| **Positive urgency** | 7.50 (2.27) | 6.89 (2.91) | 7.00 (2.57) | 7.47 (2.95) | 0.41 | .74 |
| **Lack of premeditation** | 8.22 (2.58) | 9.15 (3.11) | 9.12 (2.86) | 8.67 (2.80) | 0.86 | .47 |
| **Lack of perseverance** | 8.97 (3.08) | 8.85 (3.62) | 8.92 (3.33) | 8.65 (3.75) | 0.06 | .98 |
| **Sensation seeking** | 6.36 (2.91) | 6.78 (2.91) | 4.40 (2.94) | 6.09 (3.39) | 3.04 | .03 |

Values are means (SD).
Abbreviations: AICT, Alcohol Cue Inhibitory Control Training; AUDIT, Alcohol use Disorder Identification; NICT, Neutral Inhibitory Control Training; BDI-II, Beck Depression Inventory II; CEQ, Craving Experience Questionnaire; PANAS, Positive And Negative Affect Scale; STAI-A&B, State-Trait Anxiety Inventory; UPPS-P, impulsivity scale (short version).

Participants guessed which group they were assigned to with an accuracy of 53%, which is not significantly greater than 50% according to a binomial test (z=0.54, p=0.3).

### Primary outcome analyses

At two weeks post-discharge, 70.4% of patients (n=88/125; CI = 62.4-78.4) were abstinent, and 29.6% relapsed (n=37/125; CI =21.6-37.6). Relapse rates at 1, 3, 5 months and 1 year after discharge were reported in supplemental, eFigure1.

Logistic regression with orthogonal simple contrasts showed a significant effect of tDCS at 2 weeks post-discharge (see Table 2, (1), C1), with a difference in abstinence rate of 19% between groups receiving verum tDCS or sham tDCS (verum tDCS 79.7% [51/64; 95% CI = 69.8-89.6] vs sham tDCS 60.7% [37/61; 95% CI = 48.3-73.1]). There was no significant effect on abstinence of ICT at 2 weeks post-discharge (AICT 66.1% [47/63; 95% CI = 54.2- 78] vs. NICT 74.6% [41/62; 95% CI = 63.8-85.4]) (see Table 2, (1), C2). Logistic regression with orthogonal deviation contrasts revealed a higher abstinence in the verum tDCS condition associated with AICT than the other three conditions (see Table 2, (2), C1; verum tDCS-AICT 86.1% [31/36; CI = 74.6-97.6] vs. others 64% [57/89; CI = 54-74]).

**Table 2.** Logistic regression results on relapse rate at 2-weeks post discharge.

|  |  | Contrasts |  |  |  | 2-weeks post discharge |  |  |  |
| --- | --- | --- | --- | --- | --- | --- | --- | --- | --- |
|  |  | Verum<br>tDCS-AICT<br>(A) | Verum<br>tDCS -<br>NICT<br>(B) | Sham<br>tDCS -<br>AICT<br>(C) | Sham<br>tDCS -<br>NICT<br>(D) | Estimate | SE | Odds<br>Ratio | p<br>value |
| (1)<br>Orthogonal<br>simple<br>contrasts | Intercept |  |  |  |  | 0.90 | .21 | 2.46 | <.001 |
|  | C1: Verum vs<br>Sham tDCS | 1/2 | 1/2 | -1/2 | -1/2 | 0.94 | .41 | 2.56 | .03 |
|  | C2: AICT vs<br>NICT | 1/2 | -1/2 | 1/2 | -1/2 | 0.40 | .41 | 1.49 | .33 |
|  | C3: Interaction | 1/4 | -1/4 | -1/4 | 1/4 | 1.01 | .83 | 2.75 | .22 |
| (2)<br>Orthogonal<br>deviation<br>contrasts | Intercept |  |  |  |  | 0.9 | .21 | 2.46 | <.001 |
|  | C1: A > B = C =<br>D | 1 | -1/3 | -1/3 | -1/3 | 0.34 | .15 | 1.40 | .02 |
|  | C2: B > A = C =<br>D | -1/3 | 1 | -1/3 | -1/3 | 0.11 | .14 | 1.12 | .42 |
|  | C3: C > A = B =<br>D | -1/3 | -1/3 | 1 | -1/3 | -0.03 | .13 | .97 | .84 |

Abbreviations: AICT, Alcohol Cue Inhibitory Control Training; NICT, Neutral Inhibitory Control Training; SE, Standard Error. C, Contrast.
Logistic Regressions (1) with orthogonal simple contrasts to evaluate the effect of verum vs. sham tDCS and AICT vs. NICT on relapse rate and (2) orthogonal with deviation contrasts to compare the relapse rate of verum tDCS-AICT to that of other interventions.

The clinical effect of the intervention did not persist beyond two weeks after the discharge (i.e., 1, 3, 6 months, and 1 year; for more details, see supplemental eResults, eTable 1, and eFigure 1).

### Secondary outcomes analyses

We found no evidence that the type of intervention significantly altered scores for craving, depression, negative or positive affect, verbal fluencies, working memory and inhibitory control (see Table 3). Based on these non-significant effects, we did not include these variables in the regression model.

**Table 3.**
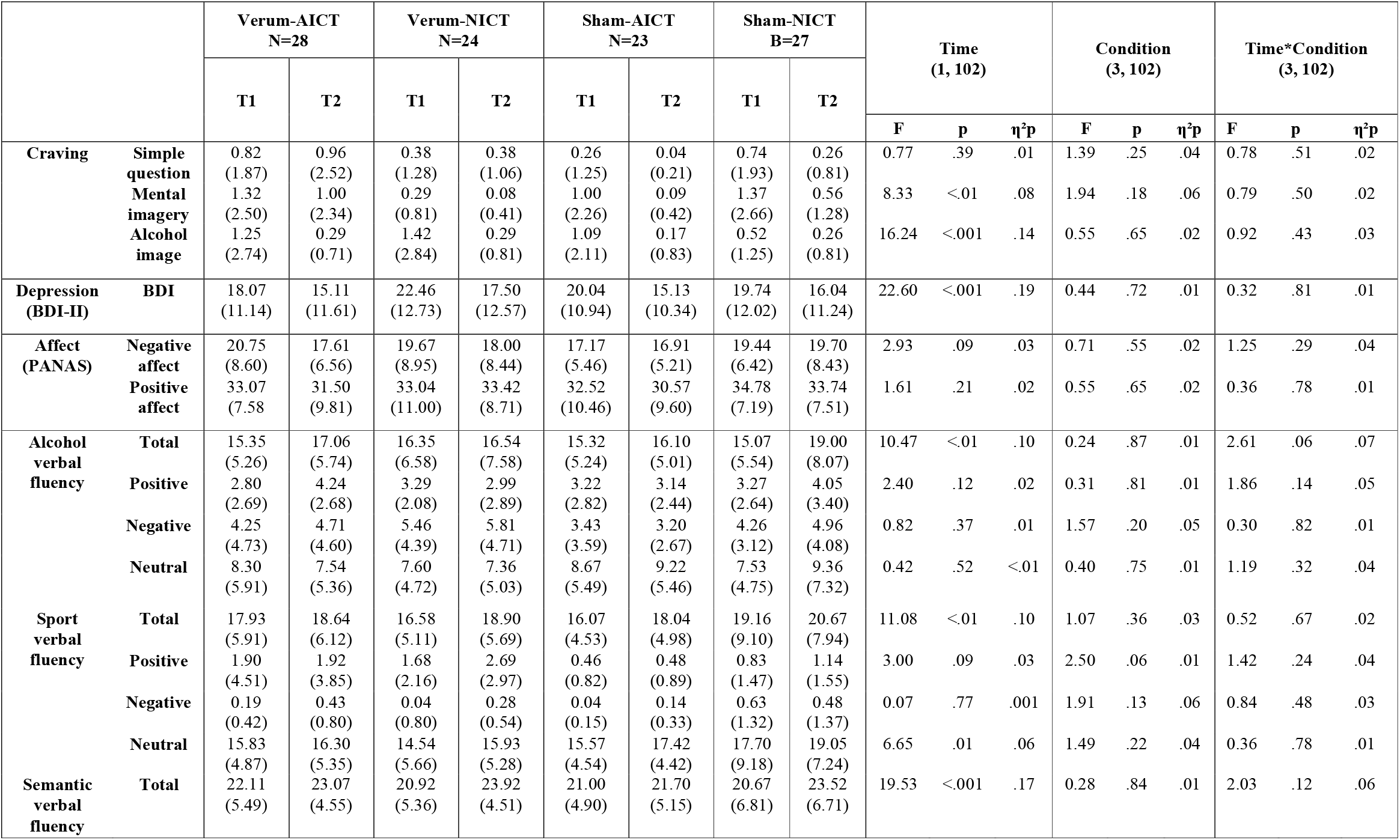
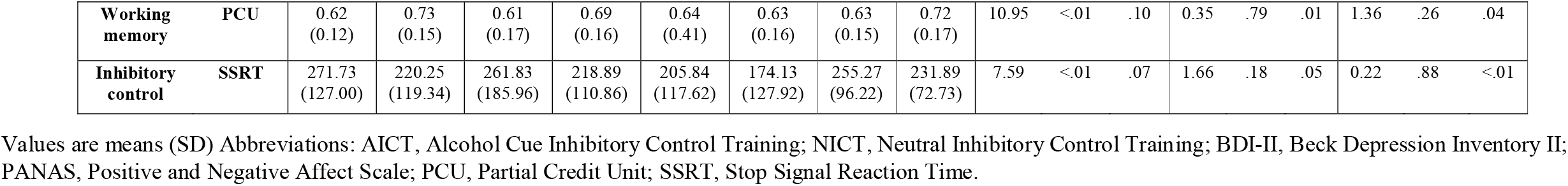
Changes in scores on craving, depression, affects, verbal fluency, working memory and inhibitory control, between baseline and after intervention.

## Discussion

The main objective of this pre-registered randomized clinical trial was to examine whether repeated sessions of tDCS applied to the DLPFC, combined with AICT, increased the likelihood of abstinence in the first two weeks following discharge. We found that regardless of the type of ICT intervention (i.e., neutral or related to alcohol), tDCS targeting the DLPFC increased the abstinence rate by 19%, compared to sham tDCS. Crucially, this tDCS intervention was not clinically beneficial when considering time points later than 2 weeks post-discharge (i.e., until one-year post-discharge). Additional analyses indicated that the combination of verum tDCS with AICT was more effective than the three other interventions. Neurocognitive interventions during alcohol rehabilitation might be efficient as an adjunctive treatment for AUD in the first weeks following discharge. This finding is very clinically relevant, as most alcohol relapses occur during this early period [3–5].

In previous studies, a positive clinical response to neurocognitive interventions was found in patients who underwent prolonged alcohol rehabilitation lasting approximately 80 days [15– 17,39,40,54,55]. However, a large proportion of detoxified participants are not willing to participate in long-term rehabilitation and are characterized by a high rate of relapse [4,5]. This observation prompted researchers to identify relevant neurocognitive interventions that protect people from alcohol relapse early after discharge [3,4,19]. Recently, a training multisession consisting of moving the image of alcohol away (by pushing a joystick) was shown to reduce early alcohol use [4]. Our findings add to the emerging literature, in that five sessions of tDCS combined with ICT aimed to improve inhibitory control in response to alcohol-related cues through associative learning was beneficial for alcohol relapse reduction during the first couple of weeks. It is noteworthy that verum tDCS improved the early abstinence rate when compared to sham tDCS, when the type of ICT was controlled for. Although the repeated practice of the alcohol-unrelated Go/No-Go cannot be confounded with the absence of cognitive training, this finding raises the possibility that multisession DLPFC stimulation could have some clinical benefits early after discharge, as recently suggested [19].

The observed synergic effects of neurostimulation and cognitive training were consistent with prior theories and recommendations [27,29,30,34]. Indeed, the “activity-selectivity” hypothesis stresses that tDCS preferentially modulates populations of active and inactive neurons [31]. Additionally, the synergistic effect found is consistent with the view that AUD results from poor inhibitory control over alcohol-related responses [7,10–12,32]. Indeed, when the prefrontal regions are stimulated, tDCS is assumed to modestly improve specific functions of executive control (e.g., response inhibition, enhanced response accuracy in online tasks, reduced aggression) [33–36,56]. On the other hand, motor response training via a specific Go/No-Go task might modify behavior by changing the explicit attraction towards an object [37,38]. However, in light of the present data, the psychological mechanisms responsible for the observed relapse protection remain unknown, as it generally does for brain stimulation and behavioral training in healthy and clinical populations [4,14,16– 18,21,40,54,55,57,58]. Indeed, we found no evidence of the intervention’s effect on potential mediators, including measures of craving, response inhibition, mood, cognitive bias and working memory.

It should be noted that a direct comparison of the present results with other trials using tDCS or ICT separately [3,4,15–17,54] or in combination [39,40], can be hazardous. Comparative studies differ in particular with regard to the number of abstinent days before the neurocognitive intervention. Nevertheless, we found support for implementing a neurocognitive intervention program in patients during the initial phase of alcohol rehabilitation.

There are several limitations to the present study. First, although the protocol was randomized, it was a single-blind study in which the participants were blinded, but the experimenters were not. Therefore, the patients could have been implicitly influenced by the experimenters’ knowledge. However, this possibility is unlikely because patients were inaccurate in guessing their assigned conditions and patients found the procedure clinically relevant to comparable degrees for all four interventions. The second limitation is the reliance on self-reported alcohol use as a key outcome instead of in-person follow-up interviews allowing biological verification of abstinence. However, the method of interview used in the present study (based on the Timeline Follow-Back method) is considered valid for measuring recent use of alcohol and other drugs [59] and has been used in a majority of studies, including a recent one similar to the present report in many aspects [4].

## Conclusion

Due to its low cost, easy availability, limited side effects, and positive impact on the clinical trajectory of the patients, we recommend the use of tDCS in association with alcohol cue ICT during alcohol rehabilitation. However, it is short-lived, and further investigations are needed to ascertain whether this intervention could be more advantageous with additional follow-up sessions after a couple of weeks. Several improvements could also be considered, including the gamification of the ICT and personalization (i.e., personal goal) of the stimuli associated with the Go and No-Go responses [60]. Finally, the identification of the psychological and neural mechanisms of the acquired resilience caused by the combination between neuromodulation and ICT remains the most intriguing question [19].

## Data Availability

the original data and analyses will be available by contacting the corresponding authors of the study as the following email address: 

## CRediT authorship contribution statement

**Macha Dubuson**: conceptualization, methodology, formal analysis, investigation, writing - original draft, supervision, project administration. **Charles Kornreich:** conceptualization, methodology, resources, writing - original draft, supervision, project administration, funding acquisition. **Marie-Anne Vanderhasselt:** conceptualization, writing - review and editing. **Chris Baeken**: conceptualization, writing - review and editing. **Florent Wyckmans:** formal analysis. **Clémence Dousset:** writing - review and editing. **Catherine Hanak:** resources. **Johannes Veeser:** resources. **Salvatore Campanella:** conceptualization, methodology, writing - review and editing. **Armand Chatard:** writing - review and editing. **Nemat Jaafari:** writing - review and editing. **Xavier Noël:** conceptualization, methodology, resources, formal analysis, writing - original draft, supervision, project administration.

## Acknowledgments

We appreciate the work of the students of the Université Libre de Bruxelles (ULB) in collecting this data, with special thanks to Sarah Sarhaoui, BA, Maaelle Brohée, MA, Tania Silva, MA, Estelle Slachmuylder, MA, Otilia-Loana Corbia, MA, and Lucie Tintlot, MA, who did not receive compensation to support this research. We thank the clinical staff of the Alcohol Unit of the CHU Brugmann for their collaboration.

## Appendix A. Supplementary data

## Appendix B. Sharing data

## Appendix C. Protocol and amendments

## Supplemental contents

### eMethods

#### Patients’ perception of the intervention

At the end of the experiment, the experimenter asked several questions to patients to elicit their opinions about various aspects of the procedure: Some examples are as follows : (1) “*Would you be prepared to continue doing this kind of computerized training for a longer period of time?”;* (2) “*Would you be willing to continue doing this cranial stimulation for a longer period of time?”;* (3) “*Do you think you were in the verum-tDCS condition?”;* (4) *“Did you find this procedure useful for your personal clinical situation?”*

#### Self-Assessment Manikin of ICT images

After the T2, the 32 images presented in the AICT (including the 16 neutral images in the NICT) were evaluated by all patients using the Self-Assessment Manikin (SAM) adapted from Bradley and Lang (1994). Patients had to respond to three Likert scales (range 1-9) for each of the 32 images: (1) Emotional valence item (negative-to-positive), with five emotional pictures ranging from distressed to happy (represented by the mouth): *“Is this image rather negative or rather positive?”;* (2) Arousal item (low to high) with five emotional pictures ranging from boredom to agitation (represented by a heart beating): *“Is this image not quite intense or stimulating, or is it rather intense or stimulating?”*. Both items were taken from Imbir 2016. Pictures were above the ratings of 1, 3, 5, 7, and 9, and there were intermediate ratings if patients felt in-between the two states; (3) Attractiveness item: *“How attractive is this image to you?’* on a 1-9 Likert scale without pictures.

#### Statistical analyses

The data during the ICT were analyzed separately from the AICT and NICT conditions because they did not have the same stimuli. We compared the first and last sessions, the correct detection, reaction time (RT), and the correct omission for neutral images between the Verum-AICT and Sham-NICT conditions. Between the Verum-AICT and Sham-AICT conditions, we compared not only the same measures but also the correct omission for alcohol images and the correct detection and RT for sports images.

We performed a one-way ANOVA on the evaluation of ICT images.

### eResults

#### Patients’ perception of the intervention

Among all patients, 76,3% reported that they would agree to continue more than five sessions of NICT and 64.5% for AICT. Among all patients, 93.1% declared they would agree to continue more than five sessions of sham-tDCS and 85.5% (91.2% in verum-AICT, 78.6% in verum-NICT, 85.2% in sham-AICT, and 100% in sham-NICT). We found no significant difference between groups on the desire to continue the two version of ICT (p=.16) and the two versions of tDCS (p=.18). To to question about the clinical utility, 88.6% of the patient found the combination verum-tDCT/AICT useful, 96.2% verum-tDCT/NICT, 95.5% sham-tDCS/AICT, 96% for sham-tDCS/NICT. No difference between groups were found (p=.55).

#### Relapse rate during one-year follow-up

At the 1-month follow-up, 119 patients were reachable; at three months, 118 patients; at six months, 115 patients; and at 1 year, 106 patients (see eFigure 1). Patients who could not be reached were classified as “relapsers”. Thus, in total, 125 patients were included in the final analysis.

**eFigure 1.**
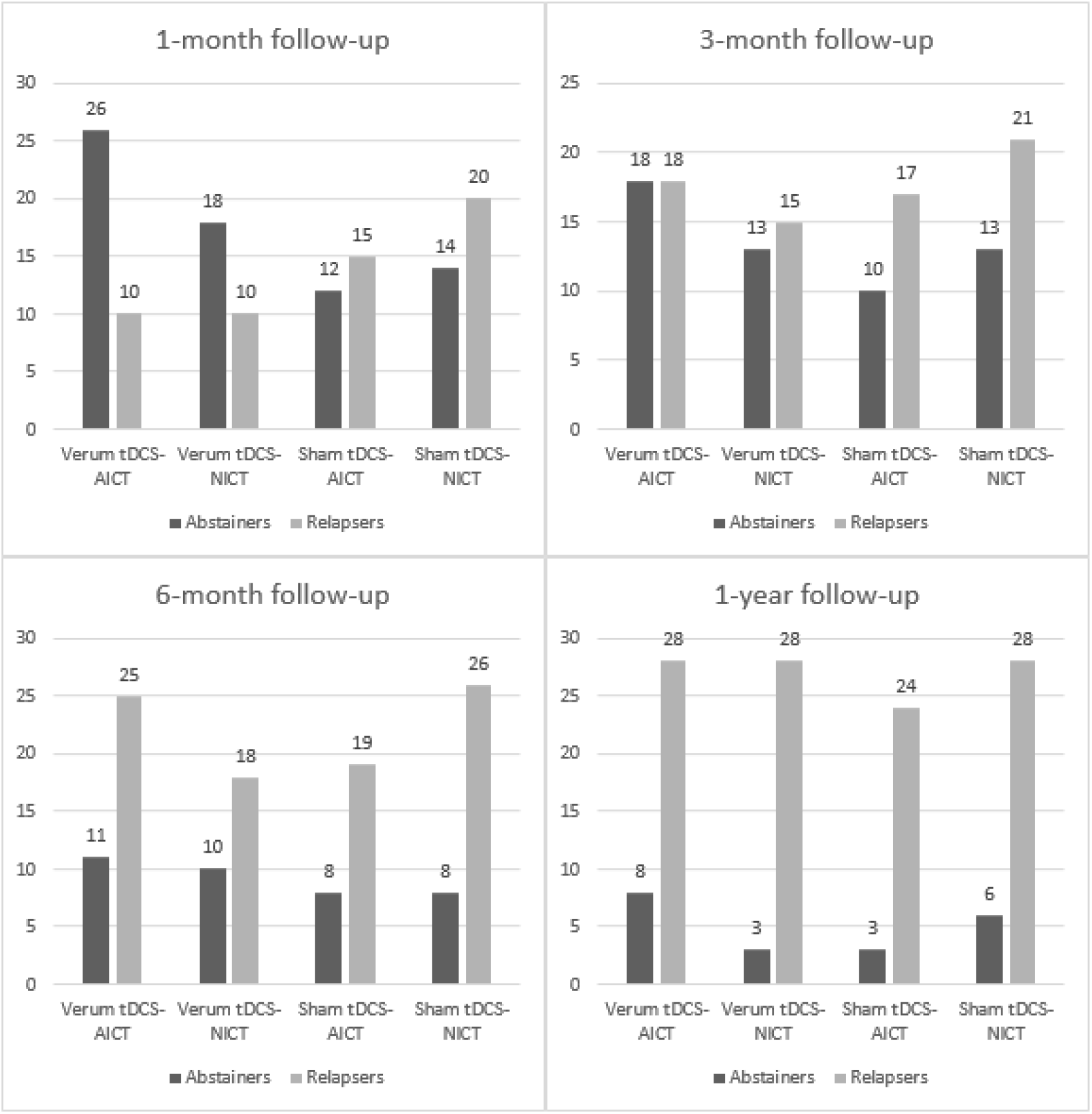
Number of relapsers and abstainers at 1-, 3-, 6-month and 1-year follow-up. Abbreviations: AICT, Alcohol cue Inhibitory Control Training; NICT, Neutral Inhibitory Control Training.

## Abbreviations

AICT: Alcohol cue Inhibitory Control Training
NICT: Neutral Inhibitory Control Training
SE: Standard Error
C: Contrast

---

Logistic Regressions (1) with orthogonal simple contrasts to evaluate the effect of verum vs. sham tDCS and AICT vs. NICT on relapse rate and (2) orthogonal with deviation contrasts to compare the relapse rate of verum tDCS-AICT to that of other interventions.

**eTable 1.** Logistic regression analyses at 1-, 3-, 6-month, and 1-year follow-up.

|  |  | 1-month follow-up |  |  |  | 3-month follow-up |  |  |  | 6-month follow-up |  |  |  | 1-year follow-up |  |  |  |
| --- | --- | --- | --- | --- | --- | --- | --- | --- | --- | --- | --- | --- | --- | --- | --- | --- | --- |
|  |  | Estimate | SE | Odds Ratio | p | Estimate | SE | Odds Ratio | p | Estimate | SE | Odds Ratio | p | Estimate | SE | Odds Ratio | p |
| (1)<br>Orthogonal<br>simple<br>contrasts | Intercept | 0.53 | 0.19 | 1.7 | < 0.01 | -0.29 | 0.18 | 0.75 | 0.12 | -0.86 | 0.2 | 0.42 | < .001 | -1.75 | 0.26 | 0.17 | < .001 |
|  | Verum vs. Sham<br>tDCS | 0.48 | 0.38 | 1.62 | 0.20 | 0.43 | 0.37 | 1.54 | 0.24 | 0.32 | 0.4 | 1.38 | 0.42 | 0.12 | 0.53 | 1.13 | 0.82 |
|  | AICT vs. NICT | 0.12 | 0.38 | 1.13 | 0.76 | 0.05 | 0.37 | 1.05 | 0.90 | 0.04 | 0.4 | 1.04 | 0.92 | 0.16 | 0.53 | 1.17 | 0.76 |
|  | Interaction | 0.5 | 0.75 | 1.65 | 0.51 | 0.19 | 0.73 | 1.21 | 0.79 | -0.55 | 0.79 | 0.58 | 0.49 | 1.41 | 1.05 | 4.1 | 0.18 |
| (2)<br>Orthogonal<br>deviation<br>contrasts | Intercept | 0.53 | 0.19 | 1.70 | < 0.01 | -0.29 | 0.18 | 0.75 | 0.12 | -0.86 | 0.2 | 0.42 | <.001 | -1.75 | 0.26 | 0.17 | <.001 |
|  | A > B = C = D | 0.15 | 0.13 | 1.16 | 0.24 | 0.12 | 0.12 | 1.13 | 0.32 | 0.09 | 0.14 | 1.09 | 0.51 | 0.07 | 0.15 | 1.07 | 0.63 |
|  | B > A = C = D | 0.06 | 0.13 | 1.06 | 0.66 | 0.08 | 0.13 | 1.08 | 0.52 | 0.15 | 0.14 | 1.16 | 0.30 | -0.14 | 0.19 | 0.87 | 0.45 |
|  | C > A = B = D | -0.03 | 0.13 | 0.97 | 0.80 | -0.01 | 0.13 | 0.99 | 0.92 | 0.08 | 0.15 | 1.08 | 0.59 | -0.13 | 0.19 | 0.88 | 0.48 |

Abbreviations: AICT, Alcohol cue Inhibitory Control Training; NICT, Neutral Inhibitory Control Training.; SE, Standard Error.

## ICTs data

Based on the AICT, six types of response measurements were calculated: correct detection RT, correct No-Go for neutral images, correct No-Go for alcohol images, and correct detection and RT for sport images. For the NICT, three scores were calculated: correct detection, RT, and correct No-Go for neutral images.

**eTable 2.**
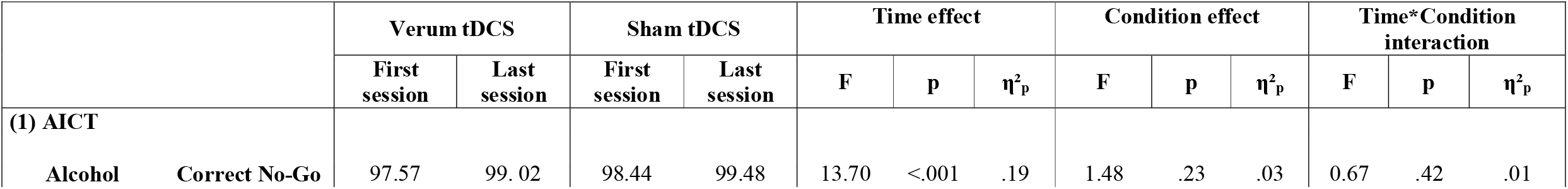
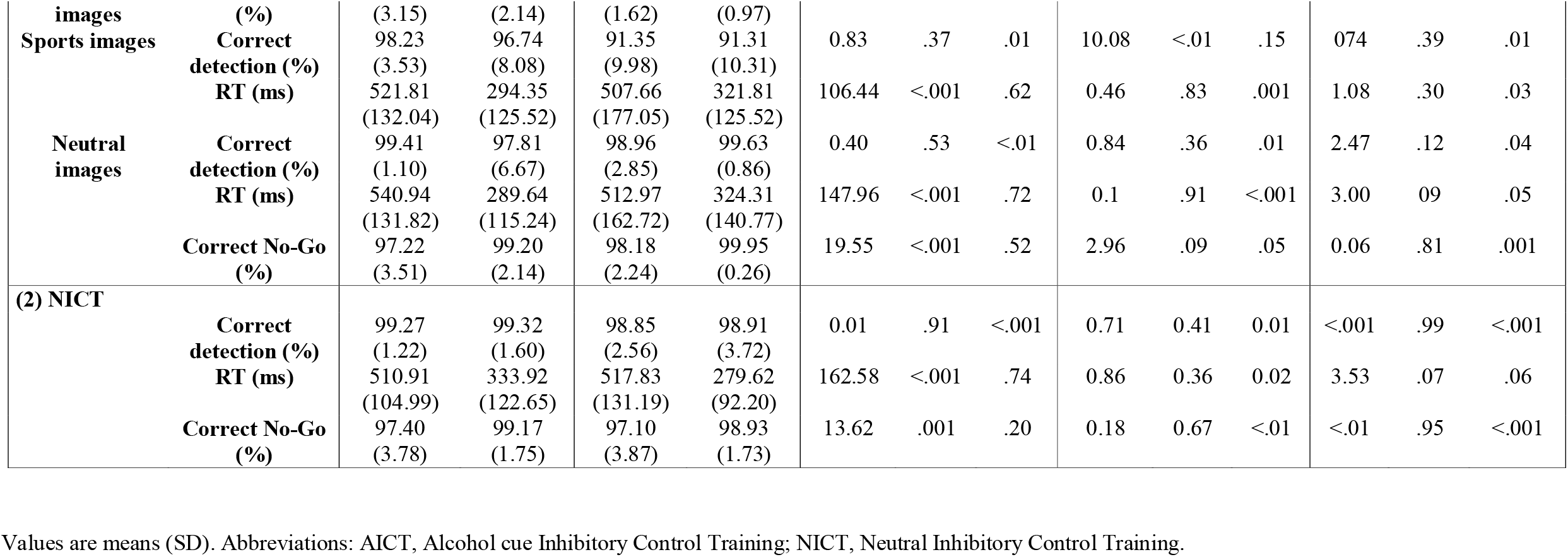
Behavioural data analyses on two versions (alcohol and neutral) of the ICT.

## Evaluation of behavioral training images

(8 alcohol images, 8 sports images, and 16 neutral images). Mean (M) and Standard Deviation (SD). One-way ANOVAs did not show any significant effect of the four conditions on Likert scores (eTable 3).

**eTable 3.** Evaluation of ICT images by the Likert scales.

| | | Verum tDCS-AICT<br>N=35 | Verum tDCS-NICT<br>N=28 | Sham-AICT<br>N=27 | Sham-NICT<br>N=32 | F<br>(3, 122) | P | $\eta^2$ |
| --- | --- | --- | --- | --- | --- | --- | --- | --- |
| **Alcohol** | **Valence** | 2.07 (1.20) | 2.15 (1.19) | 2.49 (1.52) | 2.42 (1.43) | .71 | .55 | .02 |
|  | **Arousal** | 3.19 (1.95) | 3.09 (2.51) | 3.12 (1.91) | 2.64 (1.34) | .67 | .57 | <.001 |
|  | **Attractiveness** | 3.10 (1.76) | 2.54 (1.62) | 2.78 (1.59) | 2.84 (1.48) | .64 | .59 | .009 |
| **Sport** | **Valence** | 7.86 (1.46) | 7.26 (1.64) | 8.05 (1.08) | 8.04 (1.10) | 1.83 | .15 | .04 |
|  | **Arousal** | 6.68 (1.48) | 6.98 (1.81) | 6.56 (1.74) | 7.09 (1.73) | .67 | .57 | <.01 |
|  | **Attractiveness** | 6.64 (1.48) | 7.29 (1.48) | 6.24 (1.77) | 7.10 (1.43) | 2.62 | .05 | .02 |
| **Neutral** | **Valence** | 5.68 (.83) | 5.89 (.79) | 5.38 (1.54) | 6.14 (1.43) | 2.28 | .08 | .02 |
|  | **Arousal** | 4.92 (1.35) | 5.45 (.99) | 4.53 (1.44) | 5.21 (1.44) | 2.49 | .07 | .02 |
|  | **Attractiveness** | 4.92 (1.10) | 5.29 (.96) | 4.57 (1.46) | 5.19 (1.20) | 2.07 | .11 | .02 |

Values are means (SD). Abbreviations: AICT, Alcohol-Related Inhibition Control Training; NICT, Neutral Inhibition Control Training.

